# Synthesis of High-Resolution Research-Quality MRI Data from Clinical MRI Data in Patients with COVID-19

**DOI:** 10.1101/2021.11.25.21266090

**Authors:** Ryan J. Cali, Holly J. Freeman, Benjamin Billot, Megan E. Barra, David Fischer, William R. Sanders, Susie Y. Huang, John Conklin, Bruce Fischl, Juan Eugenio Iglesias, Brian L. Edlow

## Abstract

Pathophysiological mechanisms of neurological disorders in patients with coronavirus disease 2019 (COVID-19) are poorly understood, partly because of a lack of high-resolution neuroimaging data. We applied SynthSR, a convolutional neural network that synthesizes high-resolution isotropic research-quality data from thick-slice clinical MRI data, to a cohort of 11 patients with severe COVID-19. SynthSR successfully synthesized T1-weighted MPRAGE data at 1 mm spatial resolution for all 11 patients, each of whom had at least one brain lesion. Correlations between volumetric measures derived from synthesized and acquired MPRAGE data were strong for the cortical grey matter, subcortical grey matter, brainstem, hippocampus, and hemispheric white matter (*r*=0.84 to 0.96, p≤0.001), but absent for the cerebellar white matter and corpus callosum (*r*=0.04 to 0.17, p>0.61). SynthSR creates an opportunity to quantitatively study clinical MRI scans and elucidate the pathophysiology of neurological disorders in patients with COVID-19, including those with focal lesions.

## Introduction

Neurological manifestations of coronavirus disease 2019 (COVID-19) have been recognized since the initial days of the pandemic [1]. Recent international, multi-center studies suggest that up to 80% of hospitalized patients with COVID-19 experience neurological symptoms [2], and up to 20% have altered levels of consciousness [3]. The pathogenesis of COVID-19 neurological disorders remains unclear, partly because few neuroimaging studies have been performed to investigate their associated alterations in brain structure and function [4-6]. Given the logistical challenge of enrolling patients with COVID-19 in neuroimaging studies, and the potential risk of research staff being exposed to severe acute respiratory syndrome coronavirus 2 (SARS-CoV-2), the vast majority of MRI data worldwide have been acquired for clinical purposes [7], with large slice thickness or inter-slice spacing (i.e., 5 to 6 mm) to decrease scan time (henceforth referred to as “clinical scans”). This lack of high-resolution, research-quality MRI data has hindered efforts to elucidate the pathogenesis, prognosis, and natural history of recovery from COVID-19-related neurological disorders.

We recently developed a joint super-resolution and synthesis method for generating research-quality 1 mm isotropic T1-weighted Magnetization Prepared Rapid Acquisition Gradient Echo (T1 MPRAGE) MRI data [8]. The method, termed SynthSR, involves training a convolutional neural network (CNN) with highly diverse synthetic images generated from segmentations, which makes the technique robust to changes in MR contrast, resolution, and acquisition direction of the clinical scans. SynthSR has not been tested in patients with COVID-19, nor has it been tested in patients with focal brain lesions, which are common in patients with severe COVID-19 [7]. In this retrospective study of 11 patients with neurological disorders related to severe COVID-19, we were uniquely positioned to validate SynthSR, because all patients had a clinical MRI scan that included an isotropic research-quality T1-weighted image to compare the synthesized T1 MPRAGE against. To determine the feasibility of applying SynthSR to this patient population, we tested the hypothesis that quantitative cortical and subcortical volumetric measures derived from synthesized MPRAGE images strongly correlate (*r*≥0.75) with measures derived from acquired, “gold-standard” MPRAGE images.

## Methods

### Patient Identification

We retrospectively identified patients admitted to the Neurosciences Intensive Care Unit (ICU) at our institution during the initial COVID-19 surge on the East Coast of the United States (March to June 2020) and the second COVID-19 surge (December 2020 to January 2021). Patients were included if they met the following criteria: 1) positive PCR test for SARS-CoV-2; 2) clinical diagnosis of severe COVID-19 (i.e., respiratory failure requiring mechanical ventilation); 3) clinical brain MRI performed on the 3 Tesla Skyra MRI scanner (Siemens Healthineers, Erlangen, Germany) located in our Neurosciences ICU; 4) clinical brain MRI included T2-weighted (T2) fluid-attenuated inversion recovery (FLAIR), T2, or T1 sequences (at least two of these three sequences); 5) clinical brain MRI included 1 mm isotropic T1 MPRAGE or multi-echo MPRAGE (MEMPRAGE) to provide an acquired, “gold-standard” dataset. This MRI scanner was designated for imaging patients with COVID-19 who were critically ill and therefore at high risk of having brain lesions [7]. Patients with clinical MRI performed on MRI scanners located elsewhere in the hospital were excluded to ensure consistency in the T1 MPRAGE/MEMPRAGE acquisitions and to facilitate testing SynthSR’s performance characteristics in patients with brain lesions, which has not previously been assessed [8].

### Lesion Assessments

All acquired (clinical) sequences from the clinical MRI scans were evaluated for brain lesions by two board-certified neuroradiologists (S.Y.H. and J.C.). Initial disagreements were resolved by consensus review of the images. The purpose of these evaluations was to determine if SynthSR successfully synthesizes T1 MPRAGE data in patients with lesions without overfitting (generating anatomical data that is not actually there) or underfitting (failing to produce data that otherwise should be present). We also aimed to determine whether SynthSR’s performance characteristics differ for patients with and without lesions, in terms of grey matter segmentation, white matter segmentation, and measurements of tissue volumes.

### Data Processing

To generate high-resolution synthetic images, we selected two clinical sequences with large inter-slice spacing for each subject (Supplementary Table 1). We prioritized T2 FLAIR and T1 sequences as input images for SynthSR (Figure 1A). In the absence of a T2 FLAIR or T1 sequence, we used a T2 sequence. The two clinical sequences were rigidly co-registered and entered into a 2-channel SynthSR model to generate a single, high-resolution synthesized T1 MPRAGE image for each subject. The high-resolution synthesized and acquired MPRAGEs were then processed using ‘recon-all’, a FreeSurfer function (https://surfer.nmr.mgh.harvard.edu) [9], to generate a full-brain segmentation and volumetric statistics for both datasets (Figure 1B).

**Figure 1:**
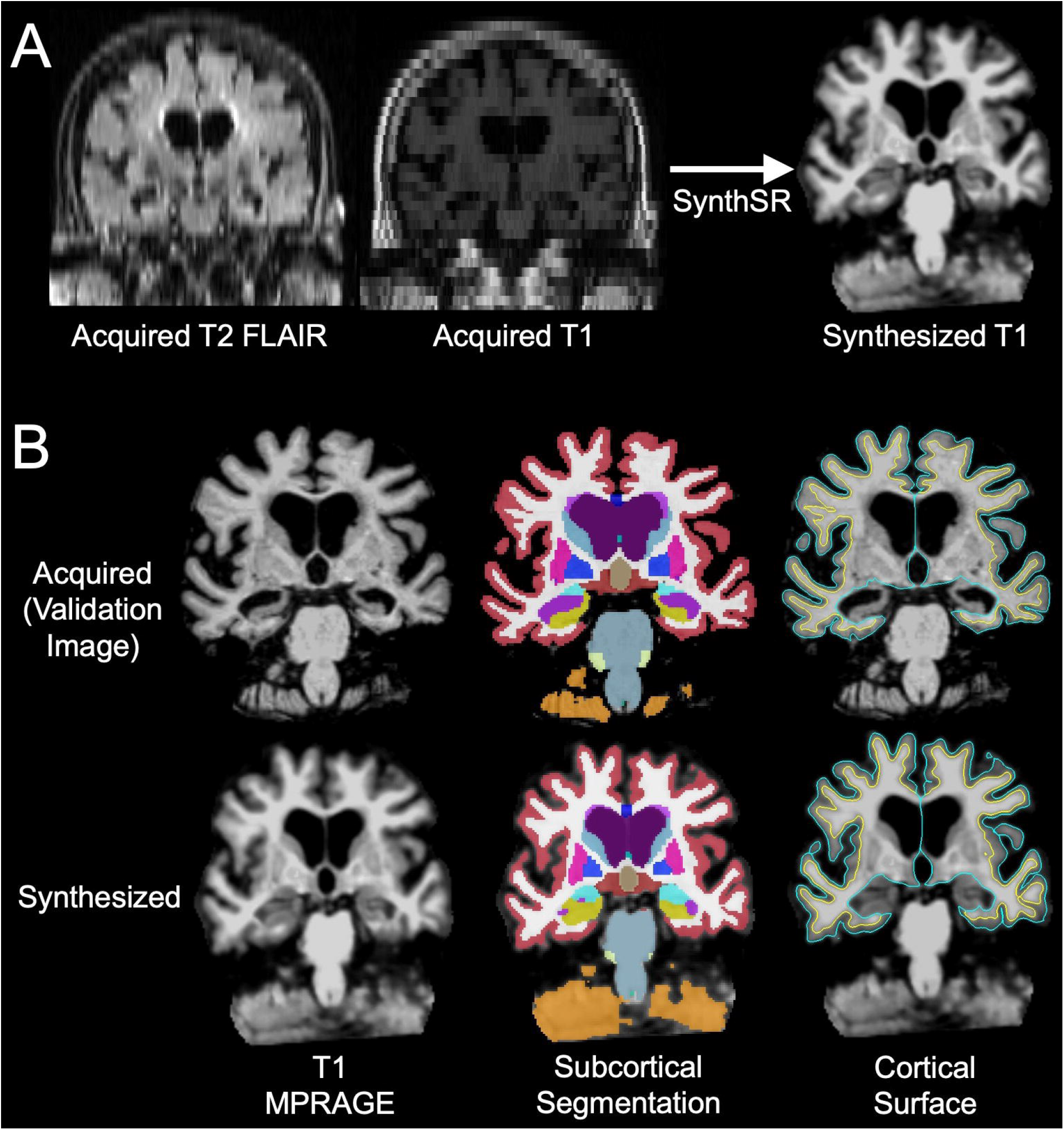
Comparison of Acquired and Synthesized Data. (A) Thick-slice T2-weighted fluid-attenuated inversion recovery (T2 FLAIR) and T1-weighted (T1) data acquired during a clinical MRI scan are used by SynthSR to synthesize a high-resolution 1mm isotropic research-quality T1 dataset. Representative data are shown from Subject 3. (B) Coronal images from subject 3 are shown from the acquired high-resolution T1 dataset (top row) and from the synthesized dataset (bottom row). Of note, each subject’s clinical MRI scan included a thick-slice T1 dataset and a 1mm isotropic T1 dataset – the latter was used to validate the synthesized dataset. The T1-weighted Magnetization Prepared Rapid Acquisition Gradient Echo (MPRAGE) MRI data are shown in the left column. FreeSurfer-based segmentation of subcortical structures is shown in the middle column, and FreeSurfer-based delineation of the cortical surface is shown in the right column. Cortical volume measurements are derived using the pial boundary (turquoise) and grey-white boundary (yellow). Consistent with the quantitative results (Table 2), this qualitative visual comparison of the acquired and synthesized datasets reveals similarities in several anatomic regions (e.g., subcortical grey matter, brainstem, cortical grey matter, and hemispheric white matter), and differences in other regions (e.g., cerebellar white matter).

### Quality Assessment (QA)

We adapted a previously published QA scale [10] to rate the anatomic accuracy of cortical surfaces and subcortical segmentations of the acquired and synthesized T1 MPRAGE datasets (Supplementary Table 2). A trained data analyst with FreeSurfer expertise (H.J.F.) performed four visual QA ratings per dataset: left and right hemisphere surface and left and right subcortical segmentation. For the synthesized datasets, we also rated whether SynthSR filled in encephalomalacic regions with inaccurate synthetic anatomy. The rater was thus not blinded to the identity of a dataset as acquired or synthesized, but the rater was blinded to the lesion ratings performed by the neuroradiologists and the cortical volumetric data.

### Statistics

We used Spearman’s Correlation Coefficient to test for associations between the acquired and synthesized volumetric measurements in 11 prespecified regions of interest. Correlations were considered significant at a Bonferroni-corrected p value <0.0045 (0.05/11).

## Results

### Patient Characteristics and Clinical MRI Findings

We identified 15 patients with severe COVID-19 who underwent a clinical MRI on the Neurosciences ICU scanner during the study period. Three were excluded because a high-resolution, isotropic T1 sequence was not acquired, and one was excluded because there were not enough clinical sequences available to generate a synthesized dataset (lacking a T2 FLAIR or T2 image). Thus, the final sample included 11 patients with severe COVID-19. There were 5 women, and the average (SD) age was 61.1 (14.7) years. Nine had lesions with diffusion restriction indicating ischemic stroke, and three had susceptibility-weighted imaging (SWI) lesions indicating intracerebral hemorrhage. Punctate SWI lesions suggestive of microhemorrhages were identified in 7, and white matter T2 hyperintensities were seen in all 11. Comprehensive demographic and clinical neuroimaging data are reported in Table 1.

**Table 1:** Demographics and Clinical Neuroimaging Characteristics.

| ID | Days from COVID Dx to MRI | Indication for MRI | Ischemic Stroke (Y/N) | If stroke present, what anatomic location? | ICH (Y/N) | If ICH present, what anatomic location? | SWI Hypointensities |  | T2 Hyperintensities |  |
| --- | --- | --- | --- | --- | --- | --- | --- | --- | --- | --- |
|  |  |  |  |  |  |  | Supra | Infra | Supra | Infra |
| 1 | 1 | L MCA syndrome | Y | L PCA | N | N/A | N | N | Y | N |
| 2 | 3 | Unresponsive | Y | L PICA, R MCA, bilateral ACA, L MCA/ACA, L MCA/PCA | N | N/A | N | N | Y | Y |
| 3 | 13 | Persistent delirium | Y | Bilateral cerebral white matter and middle cerebellar peduncles | Y | Pons, R frontal lobe, bilateral corona radiatae | Y | Y | Y | Y |
| 4 | 4 | Not following commands | Y | Splenium, R anterior temporal lobe | N | N/A | Y | N | Y | N |
| 5 | 34 | Persistently altered mental status | Y | Bilateral cerebral white matter and middle cerebellar peduncles | N | N/A | N | N | Y | Y |
| 6 | 9 | Unresponsive | N | N/A | N | N/A | N | Y | Y | N |
| 7 | 40 | Unresponsive | Y | Bilateral basal ganglia, thalami, hippocampi | N | N/A | N | N | Y | N |
| 8 | 8 | Minimal movement in extremities | Y | R ACA, R PCA, R caudate, L MCA, L splenium | Y | R medial temporal lobe, R lentiform nucleus, R gyrus rectus | Y | Y | Y | Y |
| 9 | 26 | Acute left leg weakness | Y | Bilateral frontal white matter | N | N/A | Y | Y | Y | Y |
| 10 | 0 | Unresponsive | Y | Left posterior frontal and parietal lobe | Y | L occipital lobe | Y | N | Y | N |
| 11 | 2 | Not following commands | N | N/A | N | N/A | Y | N | Y | N |

The indication for MRI was extracted from the electronic medical records. Abbreviations: ACA = anterior cerebral artery; Dx = diagnosis; ICH = intracerebral hemorrhage; Infra = infratentorial; L = left; MCA = middle cerebral artery; N = no; N/A = not applicable; PCA = posterior cerebral artery; R = right; Supra = supratentorial; SWI = susceptibility-weighted imaging; Y = yes; T2 = T2-weighted MRI.

### SynthSR Results

SynthSR successfully generated synthesized T1 MPRAGE data for all 11 patients. QA ratings revealed that for 7 of 11 patients, the quality of the FreeSurfer segmentations for the acquired and synthesized data was similar (Supplementary Table 3). For the other four patients, there were variable differences in quality of the FreeSurfer segmentations, with the acquired data quality outperforming the synthesized data quality in two, and synthesized data quality outperforming acquired data quality in two. Where differences were observed, the synthesized datasets led to more errors in subcortical segmentation, whereas the acquired datasets contained more errors in the cortical surfaces. One error unique to the synthesized data was observed in patient 2, for whom SynthSR filled in an encephalomalacic region with inaccurate anatomy (Supplementary Figure 1). Of note, this error did not prevent cortical and subcortical volumetric measures being derived in the contralateral hemisphere.

Correlations between volumetric measures derived from acquired and synthesized MPRAGE data were strong (*r* ≥ 0.75) for the subcortical grey matter (*r* = 0.96, p < 0.0001), putamen (r = 0.96, p < 0.0001), brainstem (*r* = 0.93, p < 0.0001), cortical grey matter (*r* = 0.91, p < 0.0001), hippocampus (*r* = 0.85, p = 0.0008), hemispheric white matter (*r* = 0.84, p < 0.001), globus pallidus (r = 0.83, p < 0.002), and caudate (*r* = 0.89, p = 0.0002), but non-significant (using a Bonferroni-corrected p-value threshold <0.0045) for the cerebellar cortex (*r* = 0.63, p = 0.04), and absent for the cerebellar white matter (*r* = 0.04, p = 0.91) and corpus callosum (*r* = 0.17, p = 0.61). Volumetric results for all 11 regions are provided in Table 2. Given that all 11 patients had brain lesions, it was not possible to determine if the correlation coefficients differed between patients with and without lesions.

**Table 2:** Correlations between Volumetric Measures Derived from Synthesized and Acquired T1 Volumes (n=11)

| **Neuroanatomic Region** | **Acquired T1 Volume Mean (SD)** | **Synthesized T1 Volume Mean (SD)** | **Correlation Coefficient** | **p Value** |
| --- | --- | --- | --- | --- |
| **Cortical grey matter** | 528490.5<br>(87017.4) | 558470.0<br>(70073.4) | 0.93 | <0.0001* |
| **Hemispheric white matter** | 410891.1<br>(72121.7) | 469006.6<br>(63157.0) | 0.84 | 0.001* |
| **Subcortical grey matter** | 51656.8<br>(11495.9) | 54596.0<br>(7645.7) | 0.96 | <0.0001* |
| **Cerebellar cortex** | 90378.8<br>(14084.4) | 106312.3<br>(10716.5) | 0.63 | 0.04 |
| **Cerebellar white matter** | 25293.7<br>(4796.1) | 26396.9<br>(6769.8) | 0.04 | 0.91 |
| **Corpus callosum** | 3760.4<br>(1142.2) | 2797.4<br>(391.1) | 0.17 | 0.61 |
| **Brainstem** | 19347.7<br>(3585.44) | 18233.6<br>(2527.04) | 0.93 | <0.0001* |
| **Hippocampus** | 7071.3<br>(1886.9) | 8290.4<br>(1051.2) | 0.85 | 0.0008* |
| **Putamen** | 8377.9<br>(2663.6) | 8297.5<br>(1962.6) | 0.96 | <0.0001* |
| **Caudate** | 6480.9<br>(1446.4) | 6524.5<br>(719.0) | 0.89 | 0.0002* |
| **Globus Pallidus** | 3831.2<br>(951.2) | 3529.9<br>(862.6) | 0.83 | 0.002* |

Correlations were tested using Spearman's correlation coefficient. For bilateral regions, the left- and right-sided measurements were averaged. The corpus callosum is subdivided into 5 anatomical regions within FreeSurfer. These subdivisions were summed to create the corpus callosum measurement. \* indicates a statistically significant Bonferroni-corrected p value ( $p < 0.0045$ ).

## Discussion

Using a newly developed synthesis method based on a convolutional neural network (SynthSR), we demonstrate that high-resolution research-quality MRI data can be synthesized from clinical MRI data in patients with COVID-19. In eight of 11 prespecified anatomic regions, the quantitative volumetric measurements derived from the synthesized MPRAGE dataset strongly correlated with those derived from the acquired MPRAGE dataset, suggesting that synthesized MPRAGE data can be used to investigate clinical-anatomic correlations in future studies of COVID-19-related neurological disorders. Crucially, SynthSR successfully generated 1 mm isotropic MPRAGE datasets in the presence of hemorrhagic or ischemic lesions in all 11 patients, suggesting that SynthSR may be generalizable to patients with COVID-19 who have a broad spectrum of brain lesions [7].

The potential applications of SynthSR to COVID-19-related neurological disorders are myriad. Clinical MRI scans that were previously only amenable to qualitative analysis can now be tested for associations between cortical and subcortical volumetric measures and clinical syndromes. In future work, SynthSR could also be used in patients with multiple clinical MRI scans to assess for longitudinal volumetric changes that are associated with development of chronic “long-COVID” neurological symptoms. By combining SynthSR with lesion-mapping techniques [10], it may also be possible to identify the neuroanatomic distribution of COVID-19-related brain lesions and their associated neurological syndromes. Importantly, for SynthSR to reach its full potential, its performance characteristics will need to be improved in subcortical regions such as the cerebellar white matter and cerebellar cortex.

In summary, the SynthSR tool, which we distribute to the academic community at github.com/BBillot/SynthSR and https://surfer.nmr.mgh.harvard.edu/fswiki/SynthSR, can be used to support international collaboration in the study of COVID-19-related neurological disorders. Synthesis of high-resolution images from clinical MRI data creates an opportunity for quantitative volumetric analysis of clinical MRI scans acquired in patients with COVID-19 that otherwise would have insufficient resolution for such analysis.

## Data Availability

All data produced in the present study are available upon reasonable request to the authors.

## Notes

<u>Funding</u>: This study was supported by the James S. McDonnell Foundation COVID-19 Recovery of Consciousness Consortium, the NIH Director’s Office (DP2HD101400), the NIH National Institute of Neurological Disorders and Stroke (R21NS109627, R25NS06574309, R01NS0525851, R21NS072652, R01NS070963, R01NS083534, U01NS086625, U24NS10059103, R01NS105820), NIH National Institute on Aging (R01AG070988, R56AG064027, R01AG064027, R01AG008122, R01AG016495), NIH National Institute for Biomedical Imaging and Bioengineering (P41EB015896, R01EB023281, R01EB006758, R21EB018907, R01EB019956, P41EB030006), NIH National Institute of Mental Health (R01MH123195, R01MH121885, RF1MH123195), NIH BRAIN Initiative (RF1MH123195), BRAIN Initiative Cell Census Network (U01MH117023), Alzheimer’s Research UK (ARUK-IRG2019A-003), European Research Council (Starting Grant 677697), EPSRC-funded UCL Centre for Doctoral Training in Medical Imaging (EP/L016478/1), and the Tiny Blue Dot Foundation. This work was also made possible by resources provided by Shared Instrumentation Grants 1S10RR023401, 1S10RR019307, and 1S10RR023043. Additional support was provided by the NIH Blueprint for Neuroscience Research (U01MH093765), part of the multi-institutional Human Connectome Project.

**Conflicts of interests** BF has a financial interest in CorticoMetrics, a company whose medical pursuits focus on brain imaging and measurement technologies. BF’s interests were reviewed and are managed by Massachusetts General Hospital and Partners HealthCare in accordance with their conflict-of-interest policies.

### Competing Interest Statement

Conflicts of interests: BF has a financial interest in CorticoMetrics, a company whose medical pursuits focus on brain imaging and measurement technologies. BF's interests were reviewed and are managed by Massachusetts General Hospital and Partners HealthCare in accordance with their conflict-of-interest policies.

### Funding Statement

This study was supported by the James S. McDonnell Foundation COVID-19 Recovery of Consciousness Consortium, the NIH Directors Office (DP2HD101400), the NIH National Institute of Neurological Disorders and Stroke (R21NS109627, R25NS06574309, R01NS0525851, R21NS072652, R01NS070963, R01NS083534, U01NS086625, U24NS10059103, R01NS105820), NIH National Institute on Aging (R01AG070988, R56AG064027, R01AG064027, R01AG008122, R01AG016495), NIH National Institute for Biomedical Imaging and Bioengineering (P41EB015896, R01EB023281, R01EB006758, R21EB018907, R01EB019956, P41EB030006), NIH National Institute of Mental Health (R01MH123195, R01MH121885, RF1MH123195), NIH BRAIN Initiative (RF1MH123195), BRAIN Initiative Cell Census Network (U01MH117023), Alzheimers Research UK (ARUK-IRG2019A-003), European Research Council (Starting Grant 677697), EPSRC-funded UCL Centre for Doctoral Training in Medical Imaging (EP/L016478/1), and the Tiny Blue Dot Foundation. This work was also made possible by resources provided by Shared Instrumentation Grants 1S10RR023401, 1S10RR019307, and 1S10RR023043. Additional support was provided by the NIH Blueprint for Neuroscience Research (U01MH093765), part of the multi-institutional Human Connectome Project.

### Author Declarations

Ethics approval This retrospective study was approved by the Mass General Brigham Institutional Review Board. Informed consent was waived given the retrospective nature of the study.

