## SUPPLEMENTARY MATERIAL (Fig. 1, Tables 1-3) for "Synthesis of High-Resolution Research-Quality MRI Data from Clinical MRI Data in Patients with COVID-19"

**SHORT REPORT – SUPPLEMENTARY MATERIAL**

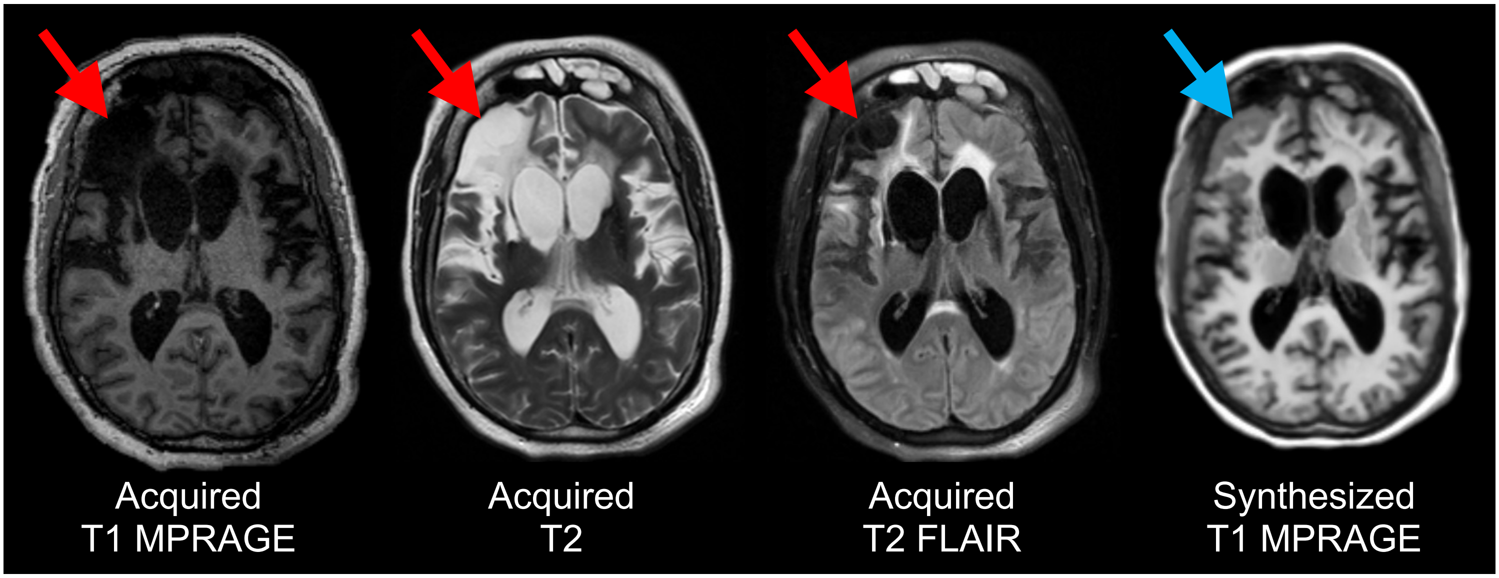


**Supplementary Figure 1: Inaccurate synthesis of brain tissue within an encephalomalacic region.** Representative axial images are shown from the acquired T1-weighted (T1) Magnetization Prepared Rapid Acquisition Gradient Echo (MPRAGE) MRI sequence, the T2-weighted (T2) MRI sequence, the T2-weighted fluid-attenuated inversion recovery (T2 FLAIR) sequence, and the synthesized T1 MPRAGE sequence from patient 2. The acquired T1 MPRAGE, T2, and T2 FLAIR images reveal a right frontal encephalomalacic lesion (red arrows; images are shown in radiologic convention). However, the synthesized T1 MPRAGE dataset shows that brain tissue has inaccurately been synthesized within this encephalomalacic region of the right frontal lobe (blue arrow).

### Supplementary Table 1: Clinical Sequence Parameters

| | | Clinical Sequence 1 | | | Clinical Sequence 2 | | |
|---|---|---|---|---|---|---|---|
| ID | Sequences (1,2) | Spatial Resolution ($mm^3$) | TE (msec) | TR (msec) | Spatial Resolution ($mm^3$) | TE (msec) | TR (msec) |
| 1 | T2 FLAIR, T1 | 0.69 x 0.69 x 6.00 | 117.00 | 4000.00 | 0.86 x 0.86 x 4.80 | 2.46 | 240.00 |
| 2 | T2 FLAIR, T2 | 0.86 x 0.86 x 6.00 | 119.00 | 9000.00 | 0.43 x 0.43 x 6.00 | 103.00 | 4940.00 |
| 3 | T2 FLAIR, T2 | 0.86 x 0.86 x 4.80 | 119.00 | 9000.00 | 0.86 x 0.86 x 4.80 | 53.00 | 2430.00 |
| 4 | T2 FLAIR, T2 | 0.94 x 0.94 x 6.00 | 119.00 | 9000.00 | 0.43 x 0.43 x 6.00 | 85.00 | 3530.00 |
| 5 | T2 FLAIR, T1 | 0.86 x 0.86 x 6.00 | 119.00 | 9000.00 | 0.86 x 0.86 x 6.00 | 53.00 | 2230.00 |
| 6 | T2 FLAIR, T2 | 0.90 x 0.90 x 6.00 | 133.00 | 9000.00 | 0.72 x 0.72 x 6.00 | 117.00 | 5500.00 |
| 7 | T2 FLAIR, T2 | 0.86 x 0.86 x 6.00 | 119.00 | 8000.00 | 0.86 x 0.86 x 4.80 | 2.46 | 240.00 |
| 8 | T2 FLAIR, T1 | 0.43 x 0.43 x 5.20 | 85.00 | 9000.00 | 0.69 x 0.69 x 5.20 | 117.00 | 5500.00 |
| 9 | T2 FLAIR, T1 | 0.86 x 0.86 x 6.00 | 119.00 | 9000.00 | 0.86 x 0.86 x 4.80 | 2.46 | 240.00 |
| 10 | T2 FLAIR, T1 | 0.90 x 0.90 x 6.00 | 119.00 | 9000.00 | 0.86 x 0.86 x 6.00 | 53.00 | 2230.00 |
| 11 | T2 FLAIR, T1 | 0.94 x 0.94 x 6.00 | 119.00 | 9000.00 | 0.75 x 0.75 x 6.00 | 2.49 | 250.00 |

Abbreviations: T2 FLAIR = T2-weighted fluid-attenuated inversion recovery; T1 = T1-weighted; T2 = T2-weighted; TE = echo time; TR = repetition time.

### Supplementary Table 2: Quality Assessment Rating Scale

| Rating | Subcortical Segmentation | Cortical Surface |
|---|---|---|
| **1** | • subcortical GM/WM segmentation is free of minor errors<br>• subcortical and cerebellar contrast is not oversaturated<br>• subcortical and cerebellar segmentations are free of minor errors and deviations | • pial and GM-WM boundary surfaces closely follow anatomical boundaries<br>• no minor areas excluded from either surface |
| **2** | • minor deviations observable in GM/WM segmentations<br>• contrast in cerebellar and subcortical areas may be difficult to assess<br>• cerebellar and subcortical segmentations appear accurate with some over- and under-labeling<br>• requires minor manual edits | • pial and GM-WM surfaces may exclude some parts of the cortex, but no major exclusions are observable<br>• requires minor manual edits |
| **3** | • obvious deviations observable in GM/WM segmentations, slightly worse than rating '2'<br>• requires major manual edits | • obvious deviations on the surfaces requiring major manual edits, but not enough to warrant exclusion |
| **4**<br>**EXCLUDE** | • major deviations observable in GM/WM segmentations<br>• little to no contrast in subcortical and cerebellar areas<br>• major deviations in subcortical and cerebellar labels | • major disruptions in pial and GM-WM surfaces<br>• excludes and/or mislabels areas of cortex |

Abbreviations: GM = grey matter; WM = white matter

**Supplementary Table 3: Quality Assessment Ratings for FreeSurfer Segmentations**

| ID | Left Subcortical Segmentation | | Right Subcortical Segmentation | | Left Cortical Surface | | Right Cortical Surface | |
|---|---|---|---|---|---|---|---|---|
| | Acq | Synth | Acq | Synth | Acq | Synth | Acq | Synth |
| **1** | 2 | 2 | 2 | 2 | 2 | 1 | 1 | 1 |
| **2** | 2 | 2 | 3 | 3 | 1 | 1 | 4 | 4 |
| **3** | 2 | 2 | 2 | 2 | 1 | 1 | 1 | 1 |
| **4** | 1 | 2 | 1 | 2 | 1 | 1 | 2 | 1 |
| **5** | 1 | 2 | 1 | 2 | 1 | 2 | 1 | 1 |
| **6** | 2 | 2 | 2 | 2 | 2 | 2 | 2 | 2 |
| **7** | 2 | 2 | 2 | 2 | 2 | 1 | 2 | 1 |
| **8** | 2 | 2 | 2 | 2 | 2 | 2 | 2 | 2 |
| **9** | 2 | 2 | 2 | 2 | 2 | 2 | 2 | 2 |
| **10** | 2 | 2 | 2 | 2 | 2 | 2 | 2 | 2 |
| **11** | 2 | 2 | 2 | 2 | 2 | 2 | 2 | 2 |

Visual quality assessment ratings are provided for the FreeSurfer segmentations of the acquired and synthesized 1 mm isotropic T1-weighted datasets. The rating scale is described in Supplementary Table 1. Boxes highlighted in grey indicate a difference in rating between the acquired and synthesized datasets. Abbreviations: Acq = acquired T1-weighted sequence; Synth = synthesized T1-weighted sequence.
